# Ultra-fast, high throughput and inexpensive detection of SARS-CoV-2 seroconversion

**DOI:** 10.1101/2021.04.13.21255396

**Authors:** Marcelo S. Conzentino, Tatielle P. C. Santos, Khaled A. Selim, Berenike Wagner, Janette T. Alford, Nelli Deobald, Nigela. M. Paula, Fabiane G. M. Rego, Dalila L. Zanette, Mateus N. Aoki, Jeanine M. Nardin, Maria C.C. Huergo, Rodrigo A. Reis, Luciano F. Huergo

**Affiliations:** Setor Litoral, UFPR Matinhos, PR, Brazil; Organismic Interactions Department, Interfaculty Institute for Microbiology and Infection Medicine, Cluster of Excellence ‘Controlling Microbes to Fight Infections’, Tübingen University, Auf der Morgenstelle 28, 72076 Tübingen, Germany; Post-Graduation Program in Pharmaceutical Sciences, UFPR, Curitiba, PR, Brazil; Instituto Carlos Chagas – FioCruz, PR, Brazil; Hospital Erasto Gaertner, Curitiba, PR, Brazil; Prefeitura Municipal de Guaratuba; Department of cell biology, UFPR, Curitiba, PR, Brazil

**Keywords:** COVID-19, magnetic ELISA, SARS-CoV-2, high throughput, magnetic beads

## Abstract

A technique allowing high throughput, fast and low-cost quantitative analysis of human IgG antibodies reacting to SARS-CoV-2 antigens will be required to understand the levels of protecting antibodies in the population raised in response to infections and/or to immunization. We described previously a fast, simple, and inexpensive Ni^2+^ magnetic bead immunoassay which allowed detection of human antibodies reacting against the SARS-CoV-2 nucleocapsid protein using a minimal amount of serum or blood. A major drawback of the previously described system was that it only processed 12 samples simultaneously. Here we describe a manually operating inexpensive 96 well plate magnetic extraction / homogenization process which allows high throughput analysis delivering results of 96 samples in chromogenic format in 12 minutes or in fluorescent ultrafast format which takes only 7 minutes. We also show that His tag antigen purification can be performed on the fly while loading antigens to the Ni^2+^ magnetic beads in a process which takes only 12 min reducing the pre analytical time and cost. Finally, we show that the magnetic bead immunoassay is antigen flexible and can be performed using either Nucleocapsid, Spike or Spike RBD. The method performed with low inter and intra assay variability using different antigens and detection modes and was able to deliver >99.5% specificity and >95% sensitivity for a cohort of 203 pre pandemic and 63 COVID-19 positive samples.

---

The COVID-19 outbreak, caused by the novel beta coronavirus SARS-CoV-2, has posed an extraordinary threat to human health with ∼3,000,000 deaths reported worldwide by Abril 2021. So far there is no effective medication to treat the disease. Despite the successful development of efficient vaccines by different laboratories, vaccination is still limited to developed countries and only a small fraction of the world’s population has been immunized^1^. COVID-19 vaccination is extremely limiting, if any, in developing countries, and social distancing is the only effective measure to reduce the spread of the disease.

Tracking COVID-19 is of key importance to understand and mitigate the spread of the disease. COVID-19 testing can be performed by a variety of methods with the most common being molecular or antigen tests which detect active SARS-CoV-2 infections. Alternatively, immunological can be employed to detect human antibodies reacting against different SARS-CoV-2 antigens^2^. One advantage of immunological COVID-19 tests is that they remain detectable for months after convalescence and thus can be used for epidemiological surveillance studies^3^. Furthermore, with the advance of COVID-19 vaccination, these immunoassays can be applied to identify the fraction of the population which developed IgG antibodies reacting against SARS-CoV-2 antigens which are likely to reflect the protection after immunization. Since there is no knowledge on how long IgG antibodies raised against SARS-CoV-2 natural infection or to vaccination will last, large populational immune surveillance studies will be necessary in near future to depict that issue^4^. Hence, the development of simple, fast and inexpensive COVID-19 immunoassays which can provide quantitative data in high throughput format to different antigens will be key to enable such large cohort studies.

We described previously a fast, simple, and inexpensive Ni^2+^ magnetic bead immunoassay which allowed detection of human antibodies reacting against the SARS-CoV-2 nucleocapsid protein using a minimal amount of serum or blood^5^. Here we show that such system is amendable to high throughput and can deliver ultrafast results in less than 7 min with different SARS-CoV-2 antigens while maintaining low analytical cost.

## Experimental Section

Human samples were collected at Hospital Erasto Gaertner in Curitiba and Secretaria Municipal de Saúde in Guaratuba and Federal University of Paraná in Matinhos. Samples for serological analysis comprised both serum and plasma-EDTA. COVID-19 positive cases were confirmed by the detection of SARS-CoV-2 RNA by real-time RT-PCR from nasopharyngeal sample swabs. The time point of sampling of serum ranged from 1 to 100 days after PCR detection. Among the 63 COVID-positive cases there were 12 convalescents including 2 asymptomatic and 10 mild non-hospitalized cases. All remaining samples collected during day 1 to 14 during the hospitalization period. The cohort of 204 negative controls consisted of pre pandemic samples collected in 2018. For the work done in Germany, pre pandemic samples were purchased from Central BioHub GmbH (Henningsdorf, Germany). COVID-19 positive samples were from convalescent donor’s post quarantine and were self-reported PCR-positive for SARS-CoV-2. The Institutional Ethics Review Board CEP/HEG (n# 31592620.4.1001.0098) and (179/2020/BO2, University Hospital Tübingen) approved this study. Informed consent was obtained from all participants in this study. All methods were performed in accordance with the relevant guidelines and regulations.

Magnetic beads-based immunoassay. The magnetic bead-based immunoassay was developed using Ni^2+^ magnetic beads as described previously^5^. The recombinant SARS-CoV-2 N-terminal 6x His-tagged Nucleocapsid protein was expressed from the pLHSarsCoV2-N plasmid using *E. coli* BL21 (λDE3) as host. The cells were growth in 100 ml LB medium at 120 rpm at 37°C to OD_600nm_ of 0.4. The incubator temperature was lowered to 16°C, after 30 min, IPTG was added to a final concentration 0.3 mM and the culture was kept at 120 rpm at 16°C over/night. Cells were collected by centrifugation at 3,000 xg for 5 min. The cell pellet was resuspended in 25 ml of buffer 1 (Tris-HCl pH 8 50 mM and KCl 100 mM). Cells were disrupted by sonication on an ice bath. The soluble fraction was recovered after centrifugation at 20,000 xg for 10 minutes and incubated for 5 min on ice with gentle mix with 10 ml of Ni^2+^ magnetic particles (Promega cat number V8550) pre equilibrated in buffer 1. The beads were washed 2x with 25 ml of buffer 1 and 2x with 25 ml of buffer 1 containing 100 mM imidazole. Beads were resuspended in 25 ml of TBST and stored in 0.8 ml aliquots at 4°C. Starting from 100 ml of *E. coli* culture, coated beads for up to 6,000 assays could be prepared and were stored at 4°C for up to up to 6 months. In the case of Spike and Spike RBD 1 mg of His-tagged antigens purified as described previously^6,7^ were incubated with 1 ml of nickel magnetic particles in 25 ml of TBST. After 5 min at room temperature with gentle mix the beads were washed with 25 ml of TBST and resuspended in 5 ml of TBST. Loaded beads were stored in 0.8 ml aliquots at 4°C.

The magnetic bead immunoassay was performed using the 96-sample format using flat bottom plates (Cralplast). The 0.8 ml aliquots of antigen loaded beads were resuspended in 11 ml of TBST containing 1% (w/v) skimmed milk and 0.1 ml were distributed on each well of a 96 well plate (Fig. 1). Four micro liters of human serum were diluted in 0.2 ml of TBST 1x skimmed milk 1% (w/v) directly on the wells of a second 96 well plate. When indicated serum was substituted for 10 µl of freshly collected capillary blood from finger puncture. The magnetic beads were transferred to the sample plate and incubated with the investigated sample for 2 min with gentle mix. The beads were captured and loaded into sequential 2x wash steps for 30 sec in 1x TBST. The beads were incubated for 2 min with 0.15 ml goat anti-human IgG-HPR (Thermo Scientific) diluted 1:1,500 in 1x TBST, following by 2x wash steps for 30 sec TBST 1x. The beads were transferred to wells with 0.15 ml of the HPR substrate TMB (Thermo Scientific) and 5 min. The total time of the procedure takes less than 12 min. When the reactions were complete the beads were removed and OD_650nm_ recorded using a TECAN M Nano plate reader (TECAN) monochromator at bandwidth 9 nm and 25 flashes. For the process operating in the fluorescent detection mode the secondary conjugated used was goat anti-human IgG-PE (Moss Inc.) diluted 1:250 in 1x TBST and beads were transferred and homogenized for 10 s to a final plate containing TBST before fluorescent reading using a TECAN M Nano plate reader (TECAN) operating at fluorescent top reading. Excitation 545 nm (bandwidth 9 nm and 25 flashes) and emission 578 nm (bandwidth 20 nm, integration time 20 µs and Z-position at 20,000 µm).

**Figure 1.**
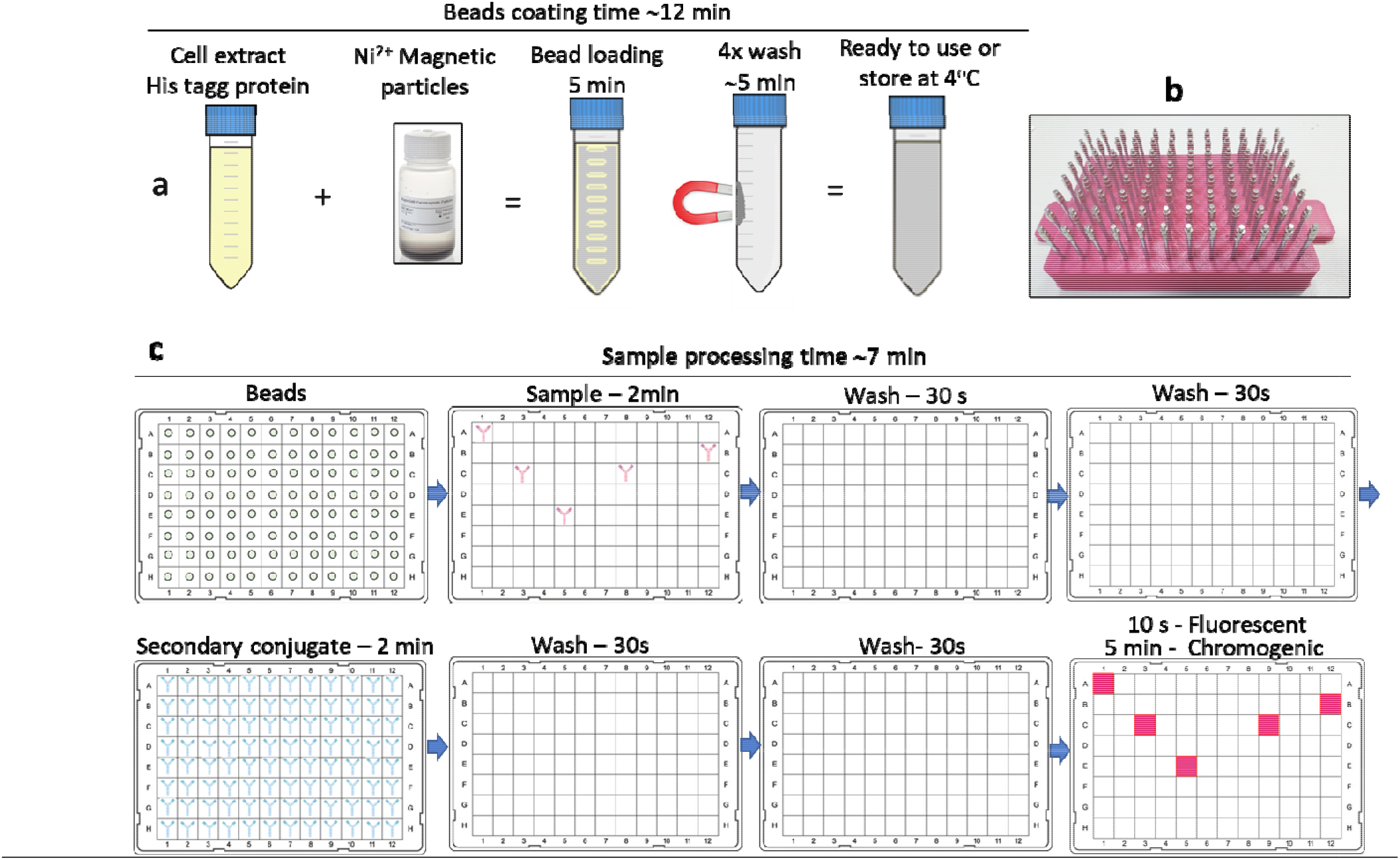
Steps of the high throughput ultrafast Ni^2+^ magnetic bead immunoassay. a) Diagram of the resin coating / antigen purification process. The soluble crude cell extract containing the 6x His tag Nucleocapsid protein is incubated with commercial Ni^2+^ magnetic particles for 5 min, followed by 4 washes with increasing imidazole concentrations. After ∼ 12 min the beads are ready to use or to store at 4°C. b) Photograph of the 96-pin magnetic extractor device used for 96-well plate magnetic bead transfer. c) Diagram of the magnetic immunoassay 8 steps process. Bead transfer from plate to plate is performed using the magnetic extractor device followed by manual homogenization for the indicated time. For fluoresce assay the final plate contains TBST buffer and just a quick 10s bead homogenization is required before fluorescent reading. For chromogenic detection, the final plate contains HPR chromogenic substrate TMB and 5 min incubation is required prior to optical density reading. Total time for fluorescent assay is ∼ 7 min.

Magnetic bead extractor device. Details of the construction of the magnetic extractor device and operation is provided in supplementary information and supplementary video.

Data analysis. One COVID-19 positive serum as reference throughout the study. Raw data were normalized as % of this reference before applying Receiver Operating Analysis (ROC) using GraphPad Prism 7.0. Statistical analysis was performed using the t test on GraphPad Prism 7.0.

## Results and Discussion

We have described previously a magnetic particle immunoassay which was successfully applied to track SARS-CoV-2 seroconversion in humans^5^. The assay principle is based on the use of commercially available Ni^2+^ magnetic particles which were coated with purified 6x His-tagged SARS-CoV-2 Nucleocapsid protein. The coated beads were used as support for a process resembling an indirect ELISA. Beads are incubated with serum or blood, washed, incubated with anti-human IgG-HPR, washed, and finally immersed on TMB, a chromogenic HPR substrate. The whole process is performed in 12 min. This immunoassay requires extraction and homogenization of the magnetic beads in different solutions. A major drawback of the system described previously was the fact that it only processes 12 sample at a time. Here we describe a manually operating inexpensive 96 well plate magnetic extraction / homogenization process which allows high throughput immunological analysis of COVID-19 cases. A 96-sample format magnetic extractor device (Fig. 1b) was built using a plastic piece fabricated on a 3D printer (Fig. S1 and S2). Inox nails were fixed to this base and a set of neodymium magnets were manually added to the top of each nail by magnetic attraction (Fig. S1). This magnetic extractor device is connected to a set of twelve 8 x 0.2 ml tubes PCR strips allowing the capture of magnetic bead particles from the solution. The magnetic particles can be transferred to another 96 well plate assisted by a set two fiber glass sticks placed underneath the PCR strips (Fig. S3). When the beads are placed on the next 96 well plate, the magnetic extractor device is lifted and separated from the PCR strips allowing the beads to go into the solution. The operator manually holds down the PCR strips and makes gentle movements back and forwards to allow complete bead homogenization (Fig. S4 and supporting video). The process is repeated until the final incubation step.

This 96-sample format process when combined to our chromogenic magnetic bead COVID-19 immunoassay was able to significantly increase the analytical throughput. The 96-sample assay was employed to investigate the presence of IgG reactive to the SARS-CoV-2 Nucleocapsid antigen in sera from 204 pre pandemic samples and 63 COVID-19 subjects. The performance obtained resembled the numbers in our previous described using the 12-sample format. Receiver operating characteristic (ROC) analysis revealed an area under curve (AUC) of 0.996. A sensitivity of 97% could be achieved at a cost of 99.5% specificity. It is important to mention that the 96-sample format maintained high intra assay / inter assay reproducibility (Table 1).

**Table 1.** Performance of the assay in different formats. 1-AUC indicate the value obtained for the area under the ROC curve. 2-Specificity was set to 99.5% (95% CI, 97.7 −99.9%) for all assays. 3 -Intra-assay CV% data obtained running the same sample in 4 wells of the same plate. 4 -Inter-assay CV% data of same sample measured in duplicate in 3 different plates.

| Assay | AUC<sup>1</sup> | % Sensitivity<sup>2</sup> | Intra-assay CV%<sup>3</sup> | Inter-assay CV%<sup>4</sup> |
| --- | --- | --- | --- | --- |
| S Chromogenic | 0.990 | 96.8 (95% CI 89.0 - 99.6) | 1.7 | 5.8 |
| S Fluorescent | 0.993 | 95.2 (95%CI 86.7 - 99.0) | 1.2 | 2.4 |
| N Chromogenic | 0.996 | 96.8 (95%CI 89.0 - 99.6) | 1.8 | 8.9 |
| N Fluorescent | 0.991 | 95.2 (95%CI 86.7 - 99.0) | 2.9 | 7.4 |

Another important key finding to increase the analytical throughput is the fact that the SARS-CoV-2 6x His tagged Nucleocapsid antigen did not need to be column-purified prior to loading the Ni^2+^ magnetic beads. Instead, soluble *E. coli* extract containing the 6x His tagged Nucleocapsid protein was directly loaded and purified on the Ni^2+^magnetic beads. This process takes only 12 min saving time and costs for protein purification (Fig. 1a).

We envisaged that the high throughput chromogenic magnetic beads immunoassay could be easily adaptable to be used with various His tagged antigens. As a proof of concept, we mobilized a His tagged version of the full-length SARS-CoV-2 Spike protein which was expressed in eukaryotic cells. These Spike coated beads were used to determine the presence of reactive IgG in the same cohort of samples. The chromogenic analysis using Spike antigen operated with AUC 0.99 and sensitivity of 97% could be achieved at a cost of 99.5% specificity (Table 1 and Fig. 2a). Again, the levels of reproducibility were high within and between different assays (Table 1). A team of operators in Germany working independently from antigen preparation to testing were able to validate the ability of the chromogenic magnetic system using His-tagged versions of either Spike or Spike RBD as antigens to discriminate COVID-19 cases (Fig. S5a). These findings support that this simple and easy to implement magnetic particle immuno-assay may be universally used with any other His tagged antigen allowing to track not only COVID-19 cases but also to investigate other diseases.

**Fig 2.**
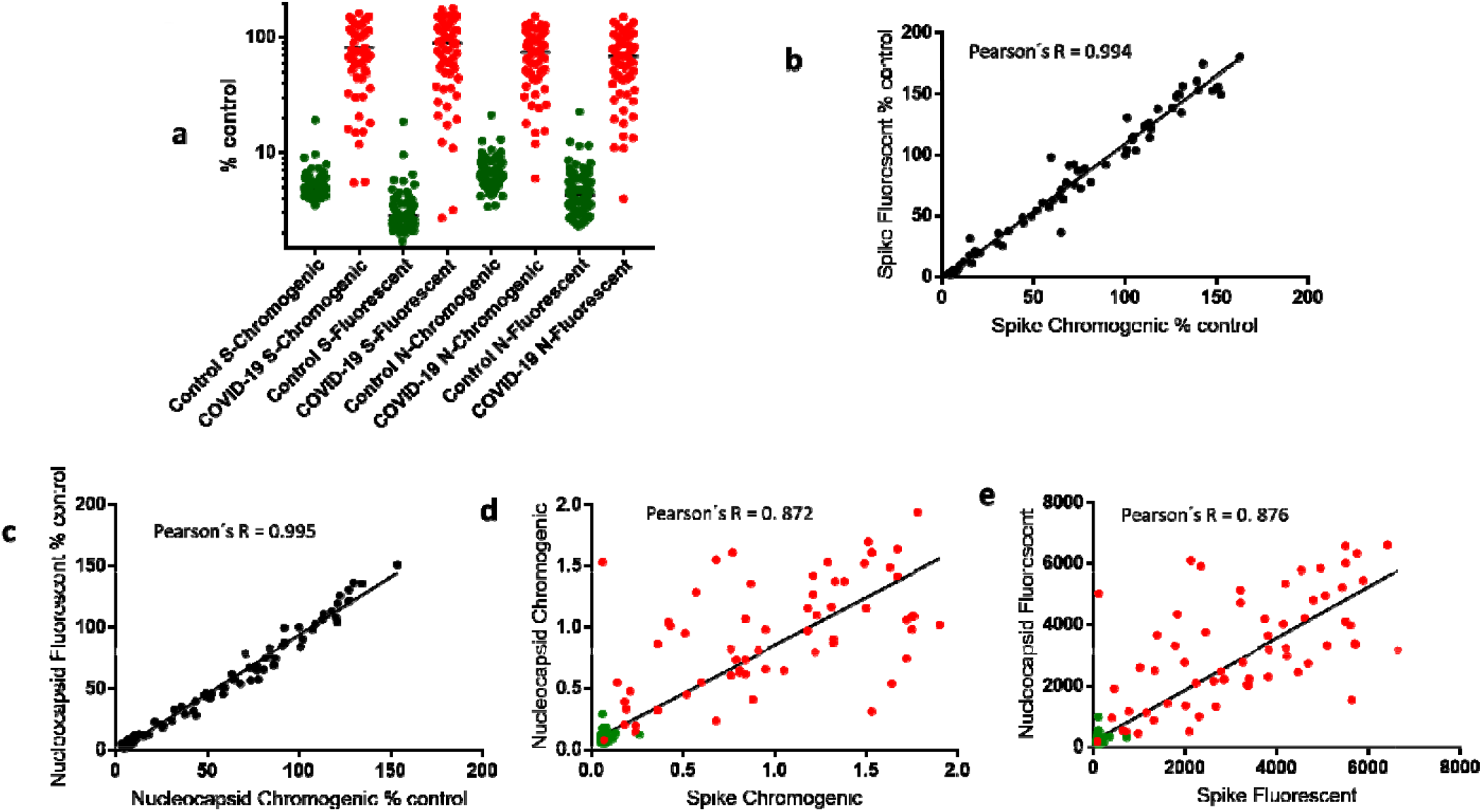
Performance of the Ni^2+^ magnetic immunoassay in different formats. a) The serum from negative controls (green) and COVID-19 positive cases (red) were analyzed using the indicate antigen (S -full length Spike or N -Nucleocapsid) and detection mode. The data was expressed as % of a calibrant control. b) Correlation between % signal obtained using Spike as antigen with Chromogenic vs fluorescent detection. c) Correlation between % signal obtained using Nucleocapsid as antigen with Chromogenic vs fluorescent detection. d) Correlation between raw signal obtained using Spike vs Nucleocapsid as antigen in chromogenic format and in fluorescent format (e).

Even though the chromogenic assay generated data in short time we envisaged that the overall reaction time could be decreased by changing the detection mode from chromogenic to fluorescent in such way that the 5 min incubation step necessary to build up oxidized TMB could be skipped. As a proof of concept, we changed the secondary anti-human IgG from HPR-conjugate to phycoerythrin-conjugate. These changes allowed an ultrafast (7 min) high throughput process which were able to discriminate COVID-19 cases using either Nucleocapsid or Spike as antigens (Fig. 2a and Table 1). The data obtained using the chromogenic and fluorescent system showed excellent correlation using serum (Fig. 2b and c) or blood (Fig. S5b). The AUC, sensitivity, specificity, and repeatability parameters of the fluorescent assay were in the same range as obtained with the chromogenic method (Table 1). The cost of the consumables per assay was down below 1$ independent of the detection mode used.

There are several debates on the literature regarding which antigen would be the best to detect COVID-19 cases with high sensitivity and specificity ^3,8,9^. Among the cohort of 204 pre-pandemic samples examined we found one sample (not the same) showing significant cross reaction for each antigen (Fig. 2a). Hence, in this study identical specificity (99.5%) was obtained using either Nucleocapsid or Spike as antigens (Table 1). The sensitivity levels were in the similar range >95% (Table 1). However, it worth mentioning that a lower background was observed in the negative cohort using Spike vs Nucleocapsid as antigen (Fig. 2a). Hence, it is likely that Spike will better perform to discriminate COVID-19 positive cases with low titer. The same argument holds true for the comparison between the detection mode. A lower background (% control) was obtained in the negative cohort using the fluorescent in comparison to chromogenic detection (Fig. 1a).

The levels of reactive IgG (raw signal) were well correlated using either Nucleocapsid or Spike as antigens (Fig. 2d and e). Two of the samples with high discrepant signal (high for Nucleocapsid and background for Spike) were collected at the day of hospitalization. Thereby, the divergent data is likely to result from ongoing activation of humoral response.

## Conclusions

Here we describe significant improvements to our Ni^2+^ magnetic beads immunoassay. Firstly, we showed that the system is amendable to high throughput analysis by employing a remarkably simple and low-cost magnetic extractor device and bead homogenization process (Fig. S1 to S4 and supporting video). Even though automated magnetic extractor and bead homogenization beads are available commercially they are unaffordable to most laboratories. Furthermore, they usually required specialized plastics (deep well plates and chip combs) which significantly increases the cost of the analysis. The process described here can be performed in regular 96-well plates and PCR tube strips (Fig. S1 to S4). The magnetic extraction / homogenization process device described may be applied to other processes such as purification of SARS-CoV-2 RNA from swab samples which routinely uses magnetic bead extraction^10^. Secondly, we show that antigen can be purified on the fly loading the cell extracts directly onto the Ni^2+^ magnetic beads in such way that 6x His tag antigen purification and bead loading occurs simultaneously in a 12 min process (Fig. 1a). Thirdly, the magnetic bead immunoassay can be performed in ultrafast 7 min format just by using an anti-human IgG PE conjugate. Fourthly, we show that the magnetic bead immunoassay can be applied using either Nucleocapsid, Spike or Spike RBD carrying a His tag as antigens (Fig. 2a). The use of Spike RBD is of particular interest as it will enables quantification of IgG with potential neutralization activity against SARS-CoV-2.

To the best of our knowledge the method described here is the only COVID-19 immunoassay which uses the principles of the well-established good standard indirect ELISA thought delivering ultrafast results in a high throughput and inexpensive format. We believe that the technique described here will be an important tool to understand the levels of immunization to previous infections and/or vaccination in large immunological surveillance studies which will be necessary to develop strategies for flexibilization of social distancing.

## Data Availability

Federal University of Parana UFPR has filed for patent protections for: the magnetic immunoassay process, magnetic COVID-19 immunological test product and magnetic bead extractor device and processing method. All designed product and processes will be freely available for academic and non-commercial users.

## Supporting Information Available

The following files are available free of charge: **Supporting information**. Details of the construction of the magnetic extractor device, operation and additional data is provided in supplementary information, figures S1 to S5. **Supporting video**. Video showing the 96-sample format magnetic bead capture and homogenization system.

## Notes

Federal University of Paraná UFPR has filed for patent protections for: the magnetic immunoassay process, magnetic COVID-19 immunological test product and magnetic bead extractor device and processing method. All designed product and processes will be freely available for academic and noncommercial users.

## Acknowledgment

We are grateful to Profa. Leda Castilho (Cell Culture Engineering Laboratory of COPPE/UFRJ) for providing the purified SARS-CoV-2 Spike protein. This work was supported by the Alexander von Humboldt foundation, UFPR, CNPq, CAPES and by the Fundação Araucária.

