## Supplementary material for "Ultra-fast, high throughput and inexpensive detection of SARS-CoV-2 seroconversion": Suplementary figures

**Supporting Information**

**Supplementary figures Legend**

**Fig S1**. **Assembly of the 96-sample magnetic extractor device**. A) Base produced in ABS plastic on a 3D printer. B) Inox nails are inserted on the plastic base using a hammer. C) Base with all 96 nails inserted. D) Final magnetic extractor device with a set of neodymium magnets added to the nails. The estimated cost of consumables for the fabrication is less than $15.

**Fig S2**. **Details of the plastic base of the magnetic extractor device**. A) Up view. B) Transsection view. Numbers indicate dimensions are in mm.

**Fig S3. Connecting the extractor device to the 96 well plate.** A) A set of 12 x 8 tubes PCR strips tubes are used. B) A flat bottom 96 well plate is filled with the magnetic beads. C) The PCR strips are placed under the wells of the plate. D and E) The magnetic extractor device is inserted into the PCR tube strips. The number 1 indicate the position of two fiber glass sticks between the strips and the 96 well plate. The number 2 indicate the position of a fiber glass stick between the PCR strips and the magnetic extractor device.

**Fig S4. Magnetic bead transfer and homogenization process**. A) The operator can lift the magnetic extractor device carrying the beads by holding the two fiber glass sticks (number 1, Fig. S2D). B) The operator can release the beads in the next solution by holding down the fiber glass stick (number 2, Fig. S2D) to keep the PCR strips on the plate while lifting the magnetic extractor device. C and D) The operator can homogenize the beads in the solution by holding down the PCR strips and making the movements back and forwards.

**Fig S5. Evaluation of the magnetic bead immunoassay using full length Spike or Spike RBD as antigens and finger puncture blood as sample.** A) The His tagged full-length Spike or the His tagged Spike RDB were mobilized in the Ni^2+^ magnetic particles and allowed to react with pre pandemic controls (green) or COVID-19 positive (red) serum using the magnetic bead system. N = 6 controls and 18 COVID-19 samples for Spike, N = 9 controls and 24 COVID-19 samples for Spike RBD. B) Ten microliters of finger puncture blood were used as sample for IgG analysis using Nucleocapsid as antigen. The % of control obtained using chromogenic vs fluorescent detection of a cohort of 61 samples are presented. Green, negative for IgG. Red, positive for IgG.

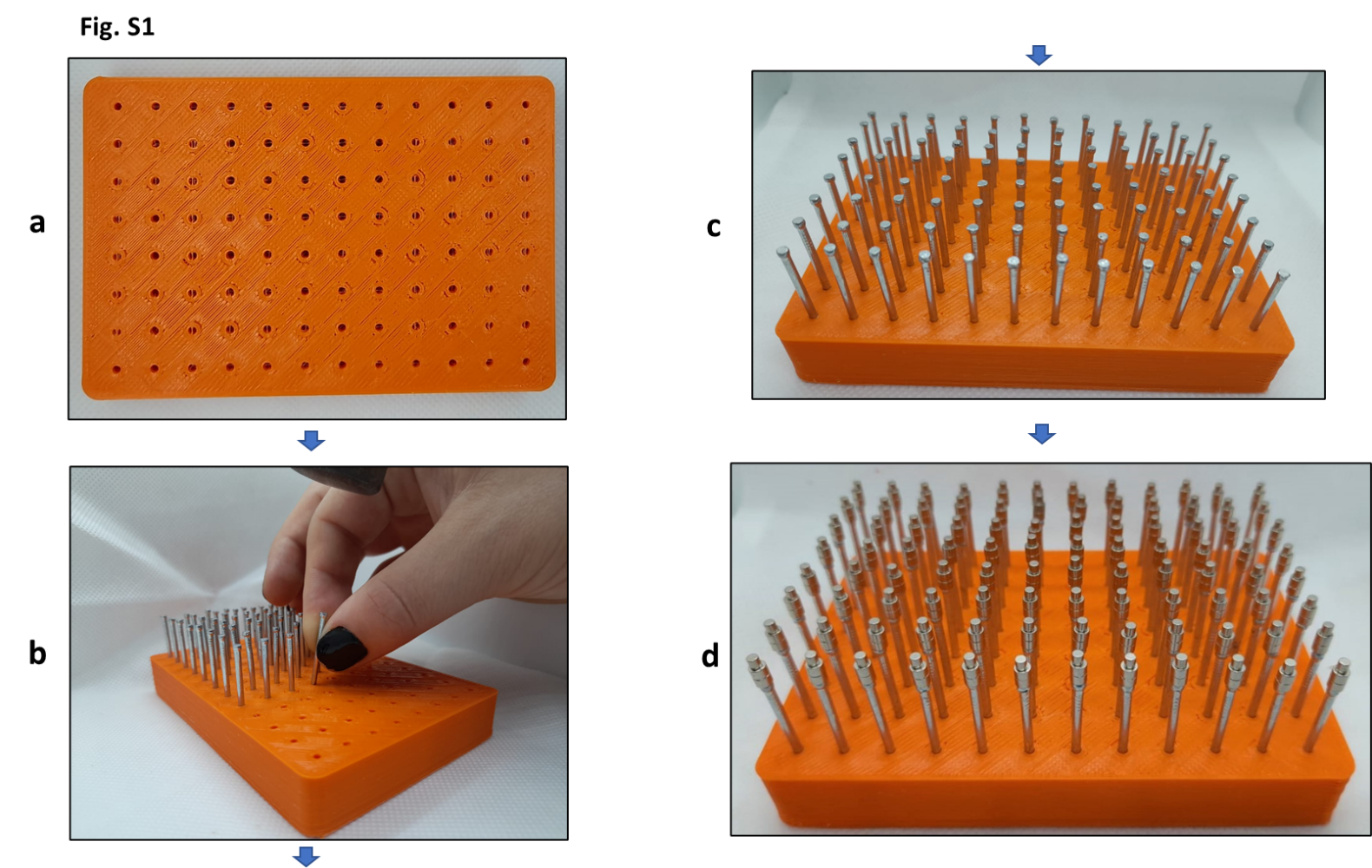

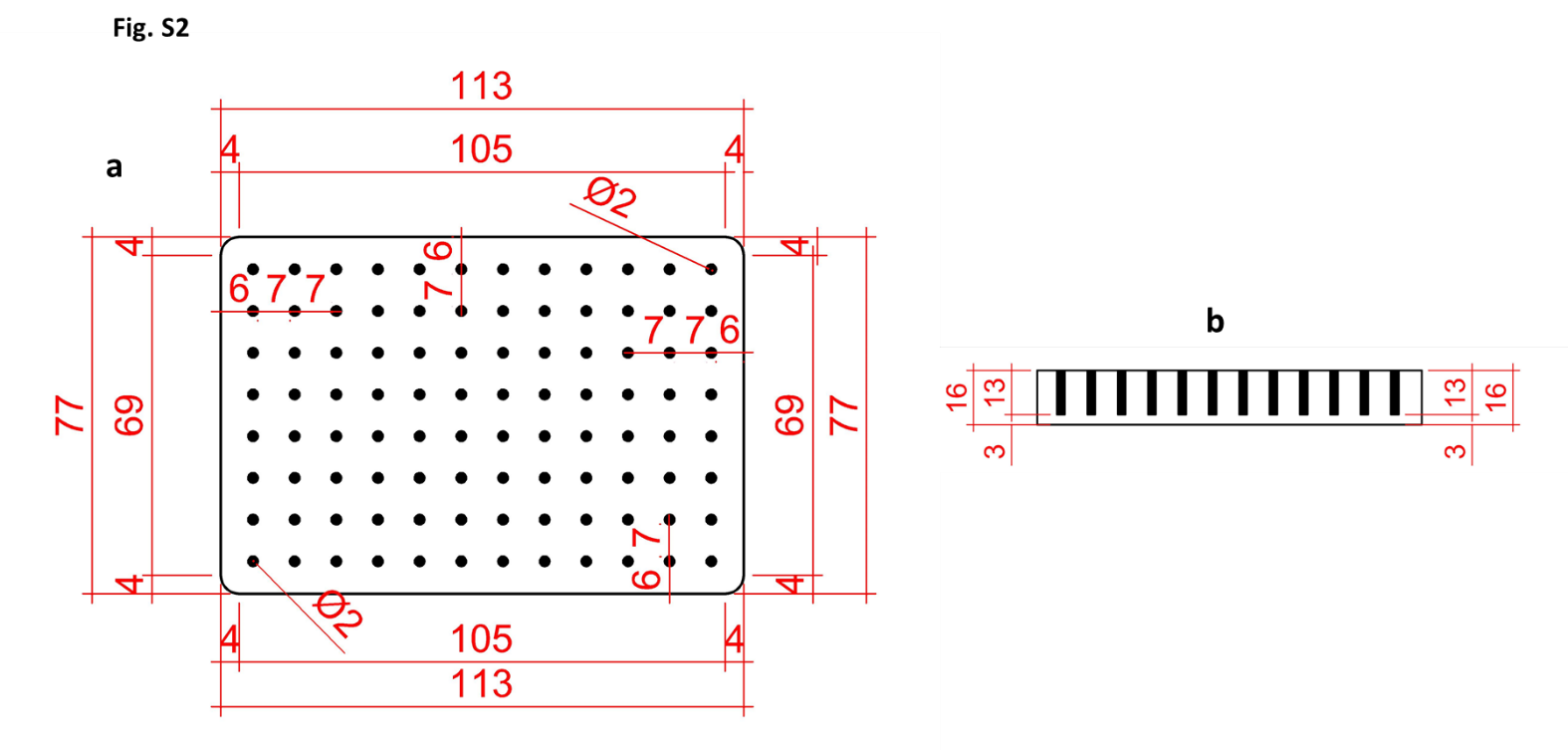

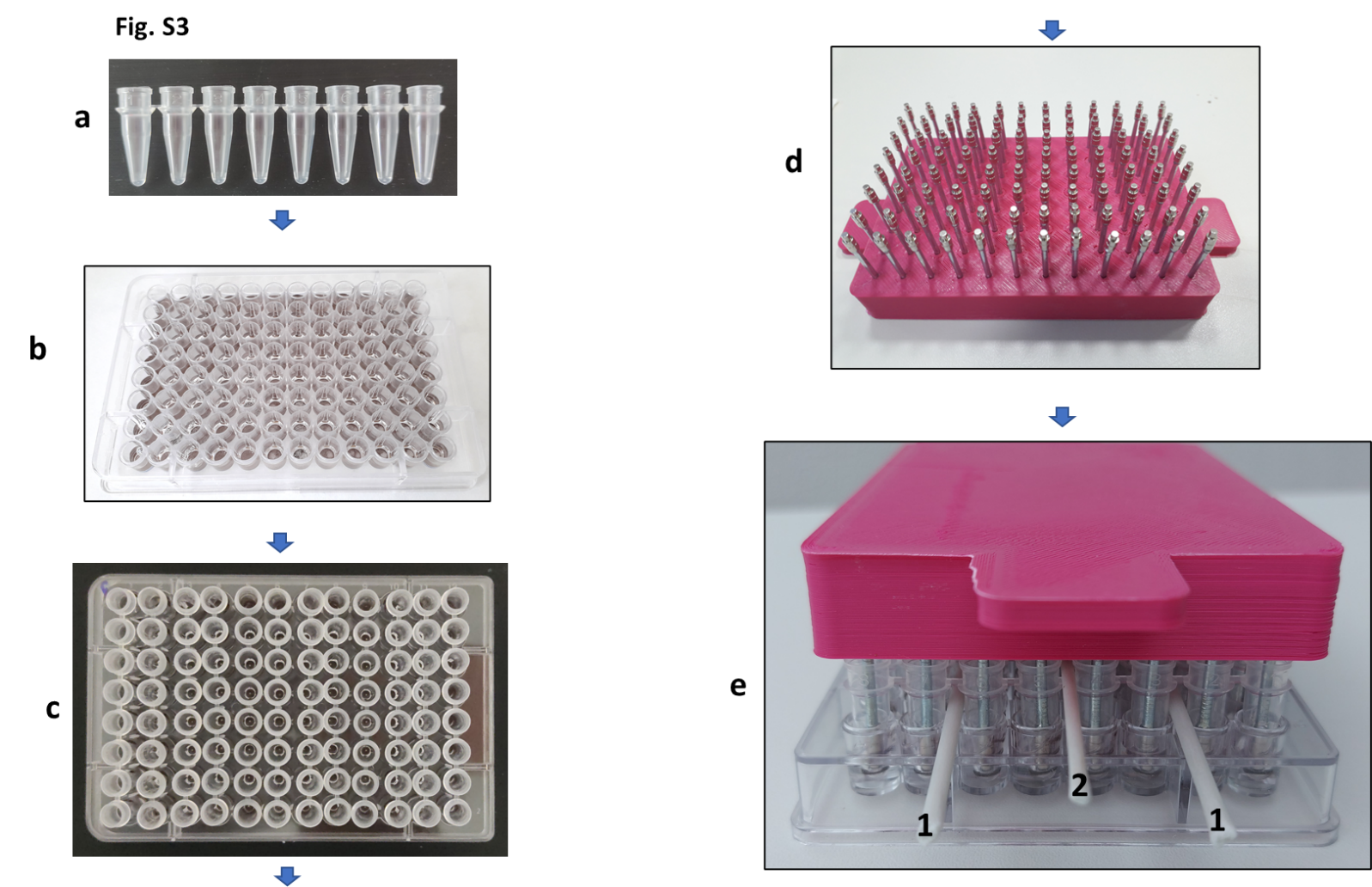

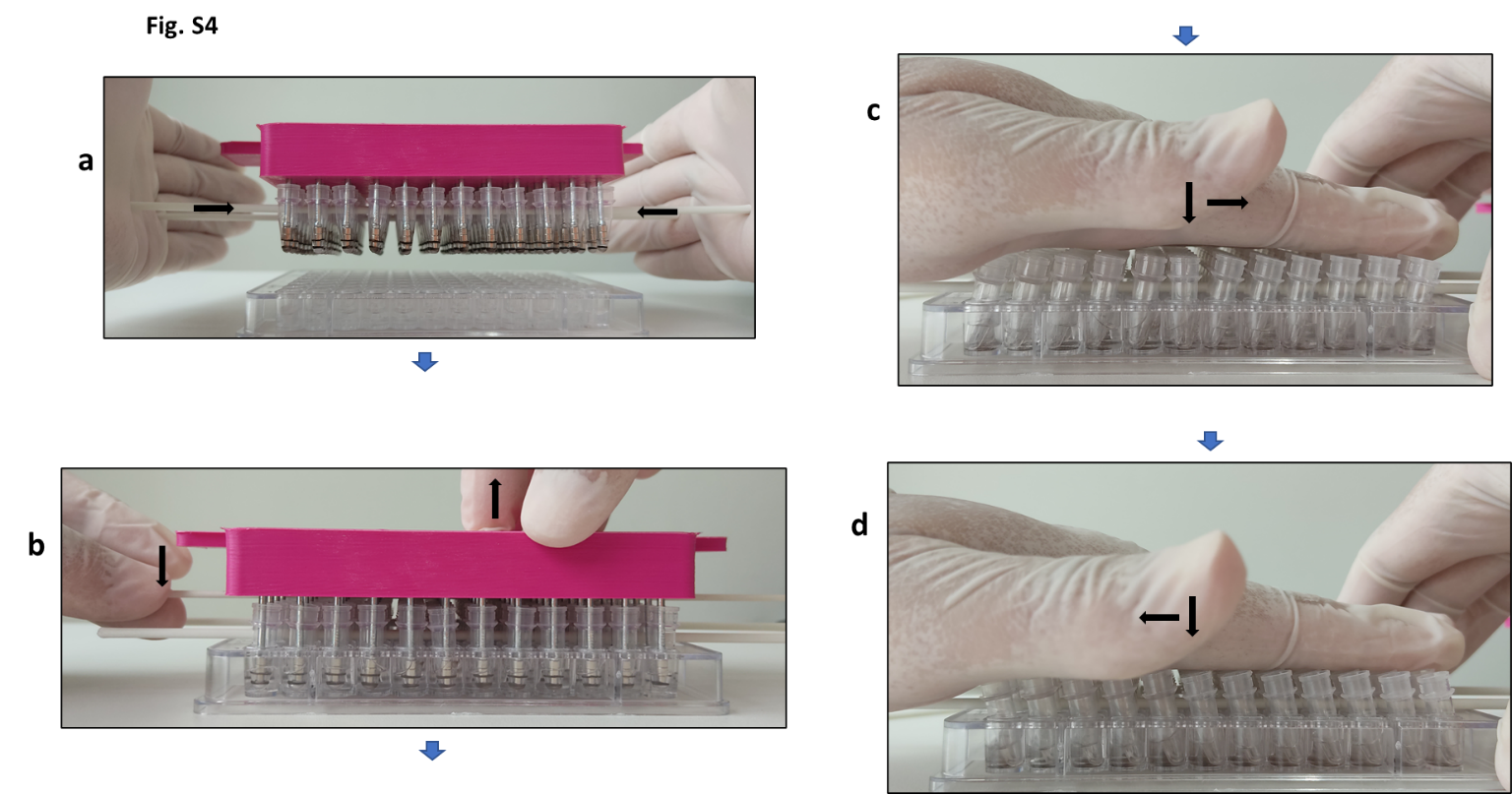

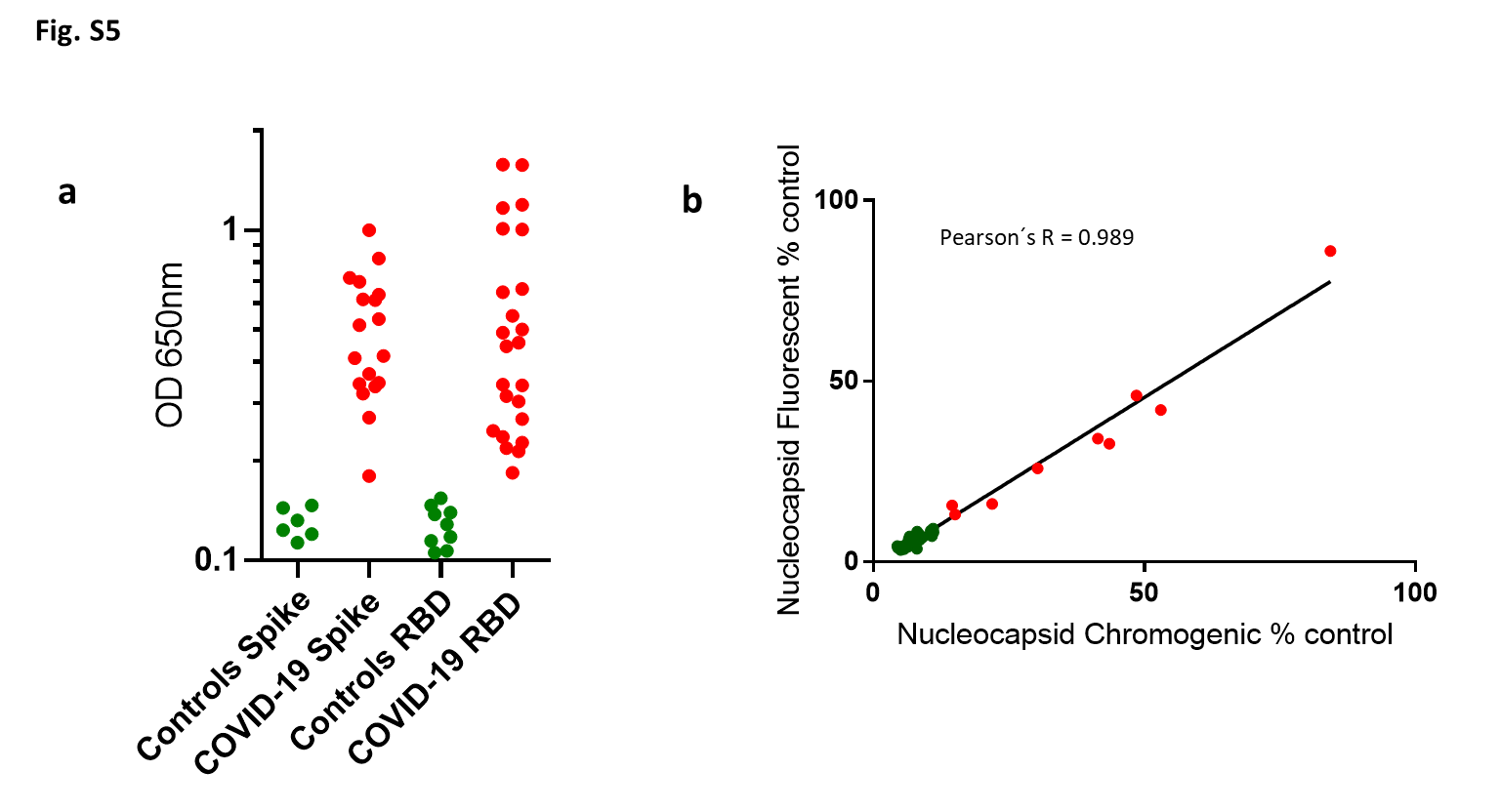
